# Prevalence and Factors Contributing to Dementia among Geriatric Patients attending Medical Outpatient Clinic at Mbeya Zonal Referral Hospital (MZRH); A Hospital Based Cross-Sectional Study

**DOI:** 10.1101/2025.08.05.25333093

**Authors:** Oscar Mahinyila, Keziah Nyaborogo

**Affiliations:** University of Dar es Salaam-Mbeya College of Health and Allied Sciences, P.O. Box 608, Mbeya, Tanzania; Department of psychiatry and Mental Health, Mbeya Zonal Referral Hospital, P.O. Box 419 Mbeya, Tanzania

**Keywords:** Dementia, Cognitive Impairment, Outpatients, Medical department, Prevalence, Factors

## Abstract

**Background:** Globally, one new case of Dementia develops in every three seconds. Patients with Cognitive Impairment usually have additional comorbidities which may accelerate progression towards a state of Cognitive Impairment. Furthermore, most suspected dementia patients visit primary physicians first instead of neurologists. This necessitates targeted screening of at-risk individuals for early intervention and prevention of Dementia.

**Aim:** This study aimed to assess the Prevalence and Factors Contributing to Dementia among Geriatric Patients attending Medical Outpatient Clinic at MZRH.

**Methods:** A cross-sectional study was conducted in April, 2024 at MZRH, in 239 patients attending medical clinic, obtained by simple random sampling. The ethical approval was obtained from UDSM-MCHAS ethics committee, which presented approval letter to MZRH administration. Identification of Dementia for Elderly Africans (IDEA) cognitive screening questionnaire was used to assess Cognitive Impairment. From a range of 0-15, the scores were grouped into three categories; ≤ 7 indicating severe Cognitive Impairment (Dementia), 8-14 indicating mild to moderate Cognitive Impairment and 15 indicating normal cognitive function. Bivariate and Multivariate analysis was performed to assess factors associated with Cognitive Impairment.

**Results:** 1 in 2 patients attending medical outpatient clinic had Cognitive Impairment. However, majority of patients had mild to moderate Cognitive Impairment (49.15%) as compared to severe Cognitive Impairment/Dementia (0.85%). Being ≥65 years old (p=0.04), lack of formal education(p=0.02), and being male (p=0.001) were significantly associated with Cognitive Impairment.

**Conclusion:** A high proportion of Cognitive Impairment was observed among elderly medical outpatients, with majority of them in early stages of Cognitive Impairment. Age, educational level, and sex of the participants significantly affected Cognitive Impairment among study participants. Cognitive screening should form part of assessment for elderly patients attending medical clinic. There is a critical need to identify new interventions that can slow progression of Cognitive Impairment advancing to Dementia, particularly in patients with additional comorbidities.

## 1.0 INTRODUCTION

### 1.1 Background

Dementia is the major cause of disability and dependency among older adults worldwide (WHO, 2017). In 2015, The World Health Organization (WHO) estimated dementia to affect 47 million people worldwide, approximately 5% of the world’s elderly population, a figure that is predicted to increase to 75 million in 2030 and 132 million by 2050. Recent reviews also estimate that globally nearly 9.9 million people develop dementia each year; this figure translates into one new case every three seconds(WHO, 2017) (Lipton & Bigal, 2016).

Dementia is an umbrella term for spectrum of diseases that progressively impair memory, behavior, and other cognitive abilities, significantly interfering with a person’s ability to maintain daily activities. Major entities in this spectrum include; Alzheimer’s disease (60-70%), vascular dementia, dementia with Lewy bodies, and a group of diseases that contribute to frontotemporal dementia. It’s a disease spectrum with indistinct entities that often co-exist (WHO, 2012).

Furthermore, a recent body of evidence suggests that, as younger age mortality is declining, the number of older people, including those living with dementia, is unexpectedly rising (Livingston et al., 2020). Nearly 60% of people with dementia currently live in low- and middle-income countries and most new cases (71%) are expected to occur in those countries (Prince, 2015; WHO, 2017). Recent data on cognitive impairment in Tanzania alerts that cognitive impairment is likely to become a significant health burden as demographic transition continues. One study conducted in rural Tanzanian population reported prevalence of dementia in this population to be similar to that reported in high-income countries(Longdon et al., 2013).

Similarly, it has been documented that the frequency of behavioral and psychological symptoms of dementia in the rural Tanzanian population to be high(S.-M. Paddick, et al., 2015) and comparable to that reported in prevalence studies from high income countries (Prince et al., 2015). Another study also reported the rates of conversion from mild cognitive impairment to dementia were similar to those reported in high-income countries (S.-M. Paddick, et al., 2015).

A study done in rural Kilimanjaro also reported prevalence of dementia was 4.6% in those aged ≥60 years and 8.9% in those aged ≥70 years. These prevalence rates increased significantly with age and also concluded that the prevalence of dementia in this rural Tanzanian population appears to have increased since 2010, although not significantly (Yoseph et al., 2021). The increasing prevalence of cognitive impairment in the general population in Sub-Saharan Africa, particularly in Tanzania, emphasizes the need for early intervention and treatment (Tai et al., 2014).

### 1.2 Problem statement

Many people with dementia have not been assessed and have not received a diagnosis. More than 95 % of those affected are over the age of 65 and the largest proportion have dementia of the Alzheimer’s type, vascular dementia or a combination of these (3). Those who are affected by other, primarily neurological diseases will often develop cognitive reduction/dementia as part of the clinical symptoms.

At present, the vast majority of dementia prevention and intervention studies are undertaken in high income countries and in white Caucasian participants (Gill Livingston et al., 2020).This implies that data on prevalence, of dementia and associated sociodemographic transitions is still inadequate in some parts of the world. Surprisingly, patients with cognitive impairment usually have additional chronic diseases (comorbidities) (Formiga et al., 2009). These chronic diseases may accelerate progression towards a state of cognitive and functional impairment leading to increased hospital visits and result in the under-diagnosis and under-treatment of dementia (Tai et al., 2014).

Conducting broad dementia screening in the general population is unreasonable because the prevalence rate of dementia and related disorders among people is too low to yield satisfactory results. However, because the risk of dementia increases substantially with age and is modified by medical commodities, the targeted screening of at-risk individuals is reasonable (Tai et al., 2014). This study aimed to assess the prevalence and associated factors of cognitive impairment and/or dementia among elderly medical outpatients at Mbeya Zonal Referral Hospital.

### 1.3 Objectives

#### 1.3.1 Broad objective

To determine the prevalence and factors contributing to dementia among geriatric patients attending Medical Outpatient Clinic at MZRH.

#### 1.3.2 Specific objectives

i. To determine prevalence of Dementia among geriatric patients attending Medical Outpatient Clinic at MZRH.
ii. To determine factors contributing to Dementia to Cognitive among geriatric patients attending medical clinic at MZRH.

### 1.4 Research questions

i. What is the prevalence of Dementia among geriatric patients attending Medical Outpatient Clinic at MZRH?
ii. What are the factors contributing to Dementia among geriatric patients attending medical Outpatient clinic at MZRH)?

### 1.5 Rationale of the study

With aging populations and evolving socio-economic structures in different parts of Tanzania, including Mbeya region, comprehending the dementia problem and its contributing factors is imperative. Therefore, findings from this study, aiming to enrich the understanding of Dementia trajectory and its connections to various contributing factors, holds the potential to inform policy decisions, addressing the this escalating health challenge.

## 3.0 METHODS

### 3.1 Study design

This was a descriptive cross-sectional study conducted in March, 2024.

### 3.2 Study area

The study was conducted at Mbeya Zonal Referral Hospital(MZRH), it is located in Mbeya region, in southern highlands of Tanzania. The hospital serves approximately 8 million people in six regions to residents from the southern highlands of Tanzania namely; Mbeya, Njombe, Iringa, Rukwa, Ruvuma and Katavi. The hospital attends a diverse range of patients with varying medical needs that are, to some extent, beyond the capacity of the primary health care settings in the zonal regions. This is carried out through its units such as Surgery, general medicine, psychiatry and mental health, pediatrics and child health, and the obstetrics, maternal health units, dental, ophthalmology and otorhinolaryngology units.

### 3.3 Study Population

The study targets elderly population aged 50 years and above, who were attending medical outpatient clinics at MZRH.

### 3.4 Sample size determination

The sample size was determined by using a Keish and Leslie formula of sample size determination as follows:

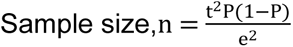

Where:

t= Standard normal variable at 95% confidence level (1.96)
e= Margin of error (0.05)
P = Existing prevalence of cognitive impairment from previous studies (18.4%) (Bickel et al., 2018).
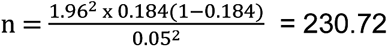, approximately 231 participants. However, 239 participants were enrolled in this study.

### 3.5 Sampling procedure

Eligible participants were selected by a simple random sampling method.

### 3.6 Inclusion and exclusion criteria

#### 3.6.1 Inclusion criteria

All inpatients aged 50 years and above, who were attending medical clinic at during the period of data collection was included in the study.

#### 3.6.2 Exclusion criteria

Patients less than 50 years, severe illness (impairing causing inability to consent, sensory impairment such as complete blindness/profound hearing, and those who were not be attending medical clinic during the period of data collection was excluded from the study.

### 3.7 Data Collection tools

The data was collected using a structured and pretested validated tool for Dementia screening adopted for African populations (S. M. Paddick et al., 2015).

#### 3.7.1 Sociodemographic characteristics

The socio-demographic characteristics such as age, sex, ability to read or write, educational level, current occupational status and the presence of chronic illnesses was collected through a structured questionnaire.

#### 3.7.2 Identification and Intervention for Dementia in Elderly Africans (IDEA) cognitive screening tool

Validated in African sites, including Tanzania. The tool was designed and validated for low literate populations, with good internal consistency and inter-rater reliability(S. M. Paddick et al., 2015). The tool is thought to perform well in comparison with other major cognitive impairment screening instruments developed for use in populations with low levels of formal education (Prince et al., 2011; Toure et al., 2012).

The tool has six items which are derived from the pre-existing cognitive assessment tools used in Low and Middle Income Countries (LMICs)(William K Gray et al., 2014). The first four items were adopted from the Community Screening instrument for Dementia (CSI-D) (Hall et al., 2000). These include being able to name a bridge from a description of its use, knowing the day of the week, knowing the name of the village chief/town mayor/ city governor and naming as many animals as possible in a minute (Scoring 2 for 8 or more animals named, score 1 for 4-7 animals named and 0 if 3 or less animals named). The fifth item was taken from Consortium to establish a Registry for Alzheimer’s Disease (CERAD) 10-word recall test (Fillenbaum et al., 2008), with a recall of 10 common words after 5 minutes’ delay (score 1 for each word up to a maximum of 5 points). The sixth item item is designed to measure praxis and involves a matchstick design test originally developed by *Baiyewu et al* (Baiyewu et al., 2005) with scores ranging from 0 (no matchsticks placed correctly), to 3 (all the four matchsticks placed correctly in shape of a rake). The maximum possible score is 15 and the minimum is 0, with a higher score indicating a better cognitive function.

Therefore, the IDEA screening tool assesses delayed recall, orientation, two measures of frontal lobe function, verbal fluency and abstract reasoning, praxis and long-term memory. An assessment of ability for new learning is also possible from the participant’s performance on the 10-word learning list. No items are included requiring reading, writing, drawing or calculation to reduce possible calculation bias.

### 3.8 Data Collection and Management

A KOBO TOOLBOX software was used in data collection, onto which structured, pre-tested questions were added to collect data. The sampled participants were informed of the purpose of the study and importance of participation and verbal consent was ensured.

### 3.9 Data processing and analysis

The data was inserted in an excel format database, and thereafter cleaned and analyzed using statistical software Stata Version 13. Data was summarized descriptively using percentages and/or proportions for categorical variables and mean and other measures of dispersion for continuous variables.

### 3.10 Ethical consideration

The permission to conduct the research was obtained from the ethical committee of University of Dar es Salaam, Mbeya College of health and allied sciences (UDSM-MCHAS) for ethical approval. After approval the letter was presented to MZRH administration. All necessary patient/participant identifiers were not known to anyone (e.g., hospital staff, patients or participants themselves) outside the research team and so cannot be used to identify individuals. The participants were informed about the objective and purpose of the study and written consent was obtained from each participant.

## 4.0 RESULTS

### 4.1 Sociodemographic characteristics of the participants

A total of 239 patients aged 50 years and above were enrolled in this study. Nearly two-third, 165 (69.04%) of the study participants were female while one-third, 74 (30.96%) were male. The mean age for participants was 66.50 years, the maximum and minimum age for study participants were 92 and 50 years respectively. After the ages of participants were grouped, the highest proportion was from 60-69 years, comprising about 92 (38.49%) of the total study participants. More than half of the study participants, 54.39% reported to have stopped working from both formal and informal occupations. Only 27 participants (11.39%) were not able to read and write. The highest education levels were in primary, about 95 (39.75%) and college and above, about 92 (38.49%). The data on sociodemographic characteristics of the study participants is presented on Table 1.

**Table 1:** Sociodemographic characteristics of the participants.

| Variable | Category | Frequency | Percentage (%) |
| --- | --- | --- | --- |
| Sex | Male | 74 | 30.96 |
|  | Female | 165 | 69.04 |
| Age groups | <65 years | 102 | 42.68 |
|  | 65 years and above | 137 | 57.32 |
| Still working? | Yes | 109 | 54.39 |
|  | No | 130 | 45.61 |
| Able to read and write | Yes | 210 | 88.61 |
|  | No | 27 | 11.39 |
| Education level | None | 27 | 11.30 |
|  | Primary school | 95 | 39.75 |
|  | Secondary school | 25 | 10.46 |
|  | College and above | 92 | 38.49 |
| Chronic illnesses | Chronic cardiovascular disease | 207 | 88.61 |
|  | Chronic lung disease | 34 | 14.23 |
|  | Chronic renal disease | 2 | 0.84 |

### 4.2 Prevalence of cognitive impairment among elderly adults attending outpatient clinics at MZRH

Out of range of 0-15 scores, participants’ scores were grouped into three categories including; ≤ 7 which implies major cognitive function impairment (dementia), 7-14 which indicates mild to moderate cognitive impairment and a score of 15 which indicates normal cognitive function. Overall prevalence of cognitive impairment among study participants was 118 (50.0%) Participants.

### 4.3 Bivariate analysis of factors associated with cognitive impairment among study participants

During data analysis, binary logistic regression was performed to investigate the relationship between cognitive function and various factors such as age, sex, education level, current occupation status, ability to read and write and presence of chronic cardiovascular, lung or renal diseases (table 2).

**Table 2.** Factors associate with Cognitive Impairment among study participants.

| **Variable** | **Prevalence of**<br>**cognitive**<br>**impairment**<br>**n (%)** | **COR**<br>**95% CI** | **P-value** | **AOR**<br>**95% CI** | **P-Value** |
| --- | --- | --- | --- | --- | --- |
| **Age** |  |  |  |  |  |
| Less than 65 years | 15 (15.0%) | Ref |  | Ref |  |
| 65 years and above | 48 (35.3%) | 3.091 | 0.001* | 2.301 | 0.040* |
| **Education level** |  |  |  |  |  |
| None | 17 (65.4%) | 11.333 | <0.001* | 22.491 | 0.020* |
| Primary school | 27 (28.7%) | 2.418 | 0.019* | 4.230 | 0.001* |
| Secondary school | 6 (24.0%) | 1.895 | 0.250 | 2.056 | 0.295 |
| College and above | 13 (14.3%) | Ref |  | Ref |  |
| **Ability to read and write** |  |  |  |  |  |
| Yes | 46 (22.2%) | Ref |  | Ref |  |
| No | 17 (63.0%) | 0.168 | <0.001* | 1.107 | 0.937 |
| **Sex** |  |  |  |  |  |
| Male | 27 (37.0%) | 2.071 | 0.018* | 3.899 | 0.001* |
| Female | 36 (22.1%) | Ref |  | Ref |  |
| **Current occupational status** |  |  |  |  |  |
| Stopped working | 38 (29.5%) | 1.370 | 0.293 | 1.094 | 0.837 |
| Still working | 25 (23.4%) | Ref |  | Ref |  |
| **Chronic cardiovascular diseases** |  |  |  |  |  |
| Yes | 52 (35.5%) | 1.618 | 0.238 | 2.222 | 0.105 |
| No | 52 (25.4%) | Ref |  | Ref |  |
| **Chronic lung diseases** |  |  |  |  |  |
| Yes | 7 (20.6%) | 1.479 | 0.387 | 1.017 | 0.991 |
| No | 56 (27.7%) | Ref |  | Ref |  |
| **Chronic renal diseases** |  |  |  |  |  |
| Yes | 0 | - |  | - | - |
| No | 63 (26.9%) | Ref |  | - | - |

\* $P < 0.05$ = statistically significant.

In a bivariate analysis, having 65 years and above, lower level of education, ability to read and write and male gender was significantly associated with cognitive impairment among study participants (p<0.05).

### 4.4 Multivariate analysis of factors associated with cognitive impairment among study participants

In a final multivariate analysis (table 2); Individuals that were aged 65 years or more were 2.3 times more likely to have cognitive impairment as compared to those below 65 years (AOR=2.301, 95% CI, (1.320-3.282), p=0.040). The chance of having cognitive impairment declined with higher education attainment, whereby individuals with no formal education were 22.5 times more likely to have cognitive impairment as compared to those with college education or higher (AOR=22.491, 95% CI, (12.425-32.557), p=0.02) and those with primary education were 4.23 times more likely to have cognitive impairment as compared to those with college education or higher (AOR=4.23, 95%CI, (2.676-5.784), p=0.001). Furthermore, male participants were 3.9 times more likely to have cognitive impairment as compared to female participants (AOR=3.899, 95% CI, (2.345-5.453), p=0.001).

## 5.0. DISCUSSION AND CONCLUSION

### 5.1 Discussion

#### 5.1.1 Prevalence of cognitive impairment among study participants

This was a hospital based cross-sectional study that aimed to estimate the prevalence of cognitive impairment among patients attending medical outpatient clinic at MZRH in the southern highlands of Tanzania. This study found that 1 in 2 patients attended at Medical outpatient clinic had cognitive impairment. However, majority of patients had mild to moderate cognitive impairment (49.15%) as compared to severe cognitive impairment (0.85%).

Epidemiological reports about cognitive impairment in elderly patients attending medical outpatient clinics from developing countries are scarce. We could not find any reported study from Sub Saharan countries reporting the prevalence of cognitive impairment in elderly by using a single-stage approach. However, most of the studies from different parts of the world such as in Malaysia (Norlaily *et al*, 2009), India (Kumari et al., 2021) and Thailand (Limpawattana et al., 2011).were done in two stage method using different screening tools or diagnostic criteria.

The prevalence of mild/moderate cognitive impairment in this study was higher compared to the prevalence reported by a study in India by Kumari et al (2021) who reported the prevalence of prevalence of mild/moderate cognitive impairment (borderline dementia) to be 20% in patients attending outpatient department of Tertiary care super specialty hospital (Kumari et al., 2021). The discrepancy may be attributed to differences in genetic/ environmental factors and screening tools employed.

The prevalence of severe cognitive impairment in this study is comparable to a study conducted in Malaysia by Norlaily *et al,* in 2009 which found the prevalence of dementia to be 2.5% among elderly patients attending outpatient clinic at University Sain Malaysi Hospital (Norlaily *et al*, 2009), using Elderly Cognitive Assessment Questionnaire (ECAQ).

Some studies however, reported higher prevalence of severe cognitive impairment (dementia) among outpatients in tertiary hospitals. These include studies conducted in Arusha, Tanzania (5%) (Roe et al., 2021), India (13.8%) (Kumari et al., 2021) and Thailand (19%) (Limpawattana et al., 2011). The discrepancy may be attributed to differences in genetic/environmental factors, assessment tools and the sample sizes employed.

Compared with healthy older adults, patients with mild cognitive impairment (MCI), accompanied by subjective cognitive decline with lack of objective cognitive and impaired learning and memory, are at a higher risk of dementia, with progression rates ranging from 16% per year (Han et al., 2022).

#### 5.1.2 Factors associated with cognitive impairment

In this study age, sex and level of education were significant factors associated with cognitive impairment.

Participants who aged 65 years or more were more likely to have cognitive impairment as compared to those aged below 65 years. These findings align with the study by Akinyemi et al which identified aging as the most significant and irreversible risk factor for stroke and cognitive impairment (Akinyemi et al., 2019).

Age is an important risk factor for cognitive decline. As we age the structure and the function of the brain deteriorates, therefore the incidence of brain atrophy is an inevitable consequence of aging(Liu-Ambrose et al., 2019). The aging process has been associated with white matter reduction in the brain, which is further correlated with reduced memory capacity (Han et al., 2022). Additionally, aging is a risk factor for extensive deterioration of multiple cognitive domains of the brain such as memory, execution, naming, attention, and reading(Han et al., 2022).

The chance of having cognitive impairment declined with higher education attainment, whereby individuals with no formal education and those with primary level of education were more likely to have cognitive impairment as compared to those with college education or higher. The findings agree with a study by Roe *et al* (2021) who reported that lacking formal education was associated with a 6 times chance of having cognitive impairment (Roe et al., 2021).

Higher levels of education are thought to be associated with increased cognitive reserve, which play a role against cognitive decline in the presence of neurodegeneration (Rosselli et al., 2022). Furthermore, the level of higher education reflects a better economic status and more extensive interpersonal communication, and the existence of these factors therefore affect the cognitive status (Kahn & Pearlin, 2006; Shin et al., 2020; Yahirun et al., 2020).

In this study, male participants were more likely to have cognitive impairment as compared to female participants. This is contrary to the findings of previous studies done in China (Han et al., 2022; Zhang, 2006) and in Lucknow-India (Kumari et al., 2021) where the prevalence of cognitive impairment was reported to be high in women than men (Kumari et al., 2021). The discrepancy can be attributed to differences in study populations, genetic or environmental characteristics in these settings. Women seem to be biologically more susceptible to Alzheimer’s disease compared to men (Podcasy & Epperson, 2016). However, this is not a law of nature, and indeed higher cognition skills are more developed in women than in men (Beam et al., 2018).

### 5.2 Conclusion

Findings from the study showed that cognitive impairment was common among patients attending medical outpatient clinic, whereby 1 in every 2 patients had cognitive impairment. However, majority of patients had mild tom moderate impairment as compared to severe cognitive impairment/dementia. Having 65 years or more, being male and low educational level were significantly associated with cognitive impairment. Furthermore, only 10.46% of the study participants had reduced performance on the instrumental activities of daily living, which also significantly associated with cognitive impairment.

## Data Availability

All relevant data are within the manuscript and its Supporting Information files.

---

**Figure 1.**
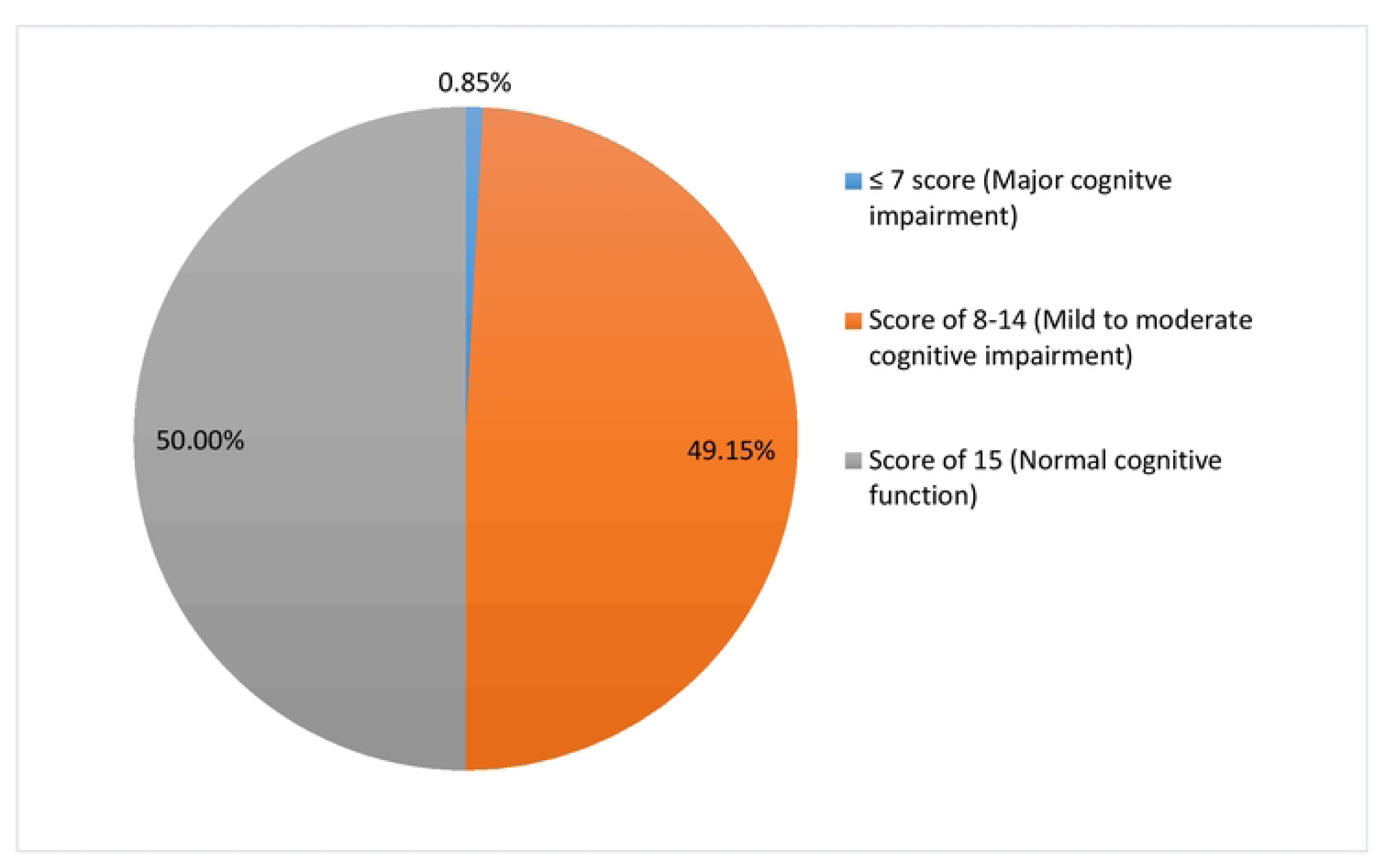
Prevalence of dementia and/or cognitive impairment among study participants.

## Notes

### Competing Interest Statement

The authors have declared no competing interest.

### Funding Statement

The author(s) received no specific funding for this work.

## References

Akinyemi, R. O., Owolabi, M. O., Ihara, M., Damasceno, A., Ogunniyi, A., Dotchin, C., Paddick, S.-M., Ogeng’o, J., Walker, R., & Kalaria, R. N. (2019). Stroke, cerebrovascular diseases and vascular cognitive impairment in Africa. Brain Research Bulletin, 145, 97–108. 10.1016/j.brainresbull.2018.05.018

American Psychiatric Association. (2022). Diagnostic and statistical Manual of Mental Disorders, Fifth Edition Text Revision (DSM-5-TR^TM^).

Baiyewu, O., Unverzagt, F. W., Lane, K. A., Gureje, O., Ogunniyi, A., Musick, B., Gao, S., Hall, K. S., & Hendrie, H. C. (2005). The Stick Design test: A new measure of visuoconstructional ability. Journal of the International Neuropsychological Society, 11(5), 598–605. 10.1017/S135561770505071X

Beam, C. R., Kaneshiro, C., Jang, J. Y., Reynolds, C. A., Pedersen, N. L., & Gatz, M. (2018). Differences Between Women and Men in Incidence Rates of Dementia and Alzheimer’s Disease. Journal of Alzheimer’s Disease, 64(4), 1077–1083. 10.3233/JAD-180141

Bickel, H., Hendlmeier, I., Heßler, J. B., Junge, M. N., Leonhardt-Achilles, S., Weber, J., & Schäufele, M. (2018). The Prevalence of Dementia and Cognitive Impairment in Hospitals. Deutsches Ärzteblatt International. 10.3238/arztebl.2018.0733

Collingwood, C., Paddick, S.-M., Kisoli, A., Dotchin, C. L., Gray, W. K., Mbowe, G., Mkenda, S., Urasa, S., Mushi, D., Chaote, P., & Walker, R. W. (2014). Development and community-based validation of the IDEA study Instrumental Activities of Daily Living (IDEA-IADL) questionnaire. Global Health Action, 7, 25988. 10.3402/gha.v7.25988

Fillenbaum, G. G., Van Belle, G., Morris, J. C., Mohs, R. C., Mirra, S. S., Davis, P. C., Tariot, P. N., Silverman, J. M., Clark, C. M., Welsh-Bohmer, K. A., & Heyman, A. (2008). Consortium to Establish a Registry for Alzheimer’s Disease (CERAD): The first twenty years. Alzheimer’s & Dementia, 4(2), 96–109. 10.1016/j.jalz.2007.08.005

Formiga, F., Fort, I., Robles, M. J., Riu, S., Sabartes, O., Barranco, E., & Catena, J. (2009). Comorbidity and clinical features in elderly patients with dementia: Differences according to dementia severity. The Journal of Nutrition, Health and Aging, 13(5), 423–427. 10.1007/s12603-009-0078-x

Gill Livingston, A. B., Jiska Cohen-Mansfield, Claudia Cooper, Sergi G Costafreda, Amit Dias, Nick Fox, Laura N Gitlin, Robert Howard, Helen C Kales, M. K., Eric B Larson, Adesola Ogunniyi, Vasiliki Orgeta, Karen Ritchie, Kenneth Rockwood, Elizabeth L Sampson, Quincy Samus, L. S. S., & Geir Selbæk, Linda Teri, N. M. (2020). Dementia prevention, intervention, and care: 2020 report of the Lancet Commission. January, 19–21.

Hall, K. S., Gao, S., Emsley, C. L., Ogunniyi, A. O., Morgan, O., & Hendrie, H. C. (2000). Community screening interview for dementia (CSI ?D?); performance in five disparate study sites. International Journal of Geriatric Psychiatry, 15(6), 521–531. 10.1002/1099-1166(200006)15:6<521::AID-GPS182>3.0.CO;2-F

Han, F., Luo, C., Lv, D., Tian, L., & Qu, C. (2022). Risk Factors Affecting Cognitive Impairment of the Elderly Aged 65 and Over: A Cross-Sectional Study. Frontiers in Aging Neuroscience, 14, 903794. 10.3389/fnagi.2022.903794

Harada, C. N., Natelson Love, M. C., & Triebel, K. L. (2013). Normal Cognitive Aging. Clinics in Geriatric Medicine, 29(4), 737–752. 10.1016/j.cger.2013.07.002

Hughes, T. M., & Sink, K. M. (2016). Hypertension and Its Role in Cognitive Function: Current Evidence and Challenges for the Future. American Journal of Hypertension, 29(2), 149–157. 10.1093/ajh/hpv180

Kahn, J. R., & Pearlin, L. I. (2006). Financial strain over the life course and health among older adults. Journal of Health and Social Behavior, 47(1), 17–31. 10.1177/002214650604700102

Kumari, R., Singh, A. K., Kaushik, A., Thakker, T., Singh, S., & Kandpal, S. D. (2021). Prevalence of dementia and its associated risk factors among elderly patient attending Outpatient Department of a tertiary care hospital in Lucknow. Indian Journal of Community Health, 33(2), 276–281. 10.47203/IJCH.2021.v33i02.009

Limpawattana, P., Sawanyawisuth, K., Soonpornrai, S., & Huangthaisong, W. (2011). Prevalence and recognition of geriatric syndromes in an outpatient clinic at a tertiary care hospital of Thailand. Asian Biomedicine, 5(4), 493–497. 10.5372/1905-7415.0504.064

Lipton, R. B., & Bigal, M. E. (2016). The epidemiology and impact of migraine. Migraine and Other Headache Disorders, May, 23–36. 10.3109/9781420019216-5

Liu-Ambrose, T., Barha, C., & Falck, R. S. (2019). Chapter Four—Active body, healthy brain: Exercise for healthy cognitive aging. In S.-Y. Yau & K.-F. So (Eds.), International Review of Neurobiology (Vol. 147, pp. 95–120). Academic Press. 10.1016/bs.irn.2019.07.004

Longdon, A. R., Paddick, S.-M., Kisoli, A., Dotchin, C., Gray, W. K., Dewhurst, F., Chaote, P., Teodorczuk, A., Dewhurst, M., Jusabani, A. M., & Walker, R. (2013). The prevalence of dementia in rural Tanzania: A cross-sectional community-based study. International Journal of Geriatric Psychiatry, 28(7), 728–737. 10.1002/gps.3880

Loy, C. T., Schofield, P. R., Turner, A. M., & Kwok, J. B. J. (2014). Genetics of dementia. The Lancet, 383(9919), 828–840. 10.1016/S0140-6736(13)60630-3

Nichols, E., Steinmetz, J. D., Vollset, S. E., Fukutaki, K., Chalek, J., Abd-Allah, F., Abdoli, A., Abualhasan, A., Abu-Gharbieh, E., Akram, T. T., Al Hamad, H., Alahdab, F., Alanezi, F. M., Alipour, V., Almustanyir, S., Amu, H., Ansari, I., Arabloo, J., Ashraf, T., … Vos, T. (2022). Estimation of the global prevalence of dementia in 2019 and forecasted prevalence in 2050: An analysis for the Global Burden of Disease Study 2019. The Lancet Public Health, 7(2), e105–e125. 10.1016/S2468-2667(21)00249-8

Norlaily, H. (2009). Proportion of Dementia and its Associated Factors Among Elderly Patients Attending Outpatient Clinics of Universiti Sains Malaysia Hospital. 64(2).

Paddick, S. M. (2015). Validation of the Identification and Intervention for Dementia in Elderly Africans (IDEA) cognitive screen in Nigeria and Tanzania. BMC Geriatrics, 15(1), 1–9. 10.1186/s12877-015-0040-1

Paddick, S.-M., Kisoli, A., Dotchin, C. L., Gray, W. K., Chaote, P., Longdon, A., & Walker, R. W. (2015). Mortality rates in community-dwelling Tanzanians with dementia and mild cognitive impairment: A 4-year follow-up study. Age and Ageing, 44(4), 636–641. 10.1093/ageing/afv048

Paddick, S.-M., Kisoli, A., Longdon, A., Dotchin, C., Gray, W. K., Chaote, P., Teodorczuk, A., & Walker, R. (2015). The prevalence and burden of behavioural and psychological symptoms of dementia in rural Tanzania. International Journal of Geriatric Psychiatry, 30(8), 815–823. 10.1002/gps.4218

Paddick, S.-M., Kisoli, A., Samuel, M., Higginson, J., Gray, W. K., Dotchin, C. L., Longdon, A. R., Teodorczuk, A., Chaote, P., & Walker, R. W. (2015). Mild Cognitive Impairment in Rural Tanzania: Prevalence, Profile, and Outcomes at 4-Year Follow-up. The American Journal of Geriatric Psychiatry : Official Journal of the American Association for Geriatric Psychiatry, 23(9), 950–959. 10.1016/j.jagp.2014.12.005

Peters, R. (2006). Ageing and the brain. Postgraduate Medical Journal, 82(964), 84–88. 10.1136/pgmj.2005.036665

Podcasy, J. L., & Epperson, C. N. (2016). Considering sex and gender in Alzheimer disease and other dementias. Dialogues in Clinical Neuroscience, 18(4), 437–446. 10.31887/DCNS.2016.18.4/cepperson

prince, martin. (2015). World Alzheimer Report.

Prince, M., Acosta, D., Ferri, C. P., Guerra, M., Huang, Y., Jacob, K. S., Llibre Rodriguez, J. J., Salas, A., Sosa, A. L., Williams, J. D., Hall, K. S., & the 10/66 Dementia Group. (2011). A brief dementia screener suitable for use by non-specialists in resource poor settings—The cross-cultural derivation and validation of the brief Community Screening Instrument for Dementia. International Journal of Geriatric Psychiatry, 26(9), 899–907. 10.1002/gps.2622

Prince, M., Wimo, A., Guerchet, M., Ali, G. C., Wu, Y.-T., & Prina, M. (2015). World Alzheimer Report 2015—The Global Impact of Dementia.

Roe, C., Safic, S., Mwaipopo, L., Dotchin, C., Klaptocz, J., Gray, K., Joseph, M., & Walker, R. (2021). 426 PREVALENCE OF, AND RISK FACTORS FOR, DEMENTIA IN ADULT OUTPATIENT REFERRALS TO A REGIONAL REFERRAL HOSPITAL IN ARUSHA, TANZANIA. Age and Ageing, 50(Supplement_2), ii5–ii7. 10.1093/ageing/afab118.01

Rosselli, M., Uribe, I. V., Ahne, E., & Shihadeh, L. (2022). Culture, Ethnicity, and Level of Education in Alzheimer’s Disease. Neurotherapeutics, 19(1), 26–54. 10.1007/s13311-022-01193-z

Shin, M., Sohn, M. K., Lee, J., Kim, D. Y., Lee, S.-G., Shin, Y.-I., Oh, G.-J., Lee, Y.-S., Joo, M. C., Han, E. Y., Han, J., Ahn, J., Chang, W. H., Shin, M. A., Choi, J. Y., Kang, S. H., Kim, Y., & Kim, Y.-H. (2020). Effect of Cognitive Reserve on Risk of Cognitive Impairment and Recovery After Stroke: The KOSCO Study. Stroke, 51(1), 99–107. 10.1161/STROKEAHA.119.026829

Tadic, M., Cuspidi, C., & Hering, D. (2016). Hypertension and cognitive dysfunction in elderly: Blood pressure management for this global burden. BMC Cardiovascular Disorders, 16(1), 208. 10.1186/s12872-016-0386-0

Tai, S.-Y., Huang, S.-W., Hsu, C.-L., Yang, C.-H., Chou, M.-C., & Yang, Y.-H. (2014). Screening Dementia in the Outpatient Department: Patients at Risk for Dementia. The Scientific World Journal, 2014, 1–6. 10.1155/2014/138786

Toure, K., Coume, M., Ndiaye, M., Zunzunegui, M. V., Bacher, Y., Diop, A. G., & Ndiaye, M. M. (2012). Risk Factors for Dementia in a Senegalese Elderly Population Aged 65 Years and Over. Dementia and Geriatric Cognitive Disorders Extra, 2(1), 160–168. 10.1159/000332022

Walker, K. A., Sharrett, A. R., Wu, A., Schneider, A. L. C., Albert, M., Lutsey, P. L., Bandeen-Roche, K., Coresh, J., Gross, A. L., Windham, B. G., Knopman, D. S., Power, M. C., Rawlings, A. M., Mosley, T. H., & Gottesman, R. F. (2019). Association of Midlife to Late-Life Blood Pressure Patterns With Incident Dementia. JAMA, 322(6), 535–545. 10.1001/jama.2019.10575

WHO. (2017). Global action plan on the public health response to dementia 2017—2025. *Geneva*: World Health Organization, 52.

William K Gray et al. (2014). Development and Validation of the Identification and Intervention for Dementia in Elderly Africans (IDEA) Study Dementia Screening Instrument. National Liobrary of Medicine. https://pubmed.ncbi.nlm.nih.gov/24578459/

World Health Organisation, WHO. (2012). Dementia: A public health priority.

Yahirun, J. J., Vasireddy, S., & Hayward, M. D. (2020). The Education of Multiple Family Members and the Life-Course Pathways to Cognitive Impairment. The Journals of Gerontology: Series B, 75(7), e113–e128. 10.1093/geronb/gbaa039

Yoseph, M., Paddick, S.-M., Gray, W. K., Andrea, D., Barber, R., Colgan, A., Dotchin, C., Urasa, S., Kisoli, A., Kissima, J., Haule, I., Rogathi, J., Safic, S., Mushi, D., Robinson, L., & Walker, R. W. (2021). Prevalence estimates of dementia in older adults in rural Kilimanjaro 2009-2010 and 2018-2019: Is there evidence of changing prevalence? International Journal of Geriatric Psychiatry, 36(6), 950–959. 10.1002/gps.5498

Zhang, Z. (2006). Gender Differentials in Cognitive Impairment and Decline of the Oldest Old in China. The Journals of Gerontology Series B: Psychological Sciences and Social Sciences, 61(2), S107–S115. 10.1093/geronb/61.2.S107

